# Exploring the needs and uses for drones in medical emergencies in England and Scotland: a survey of emergency healthcare workers

**DOI:** 10.1101/2022.10.18.22280902

**Authors:** Olivia Théorêt, Sophie Barrack, Heather May Morgan

**Affiliations:** Department of Applied Health Sciences, University of Aberdeen; The School of Medicine, Medical Sciences, and Nutrition, University of Aberdeen

**Keywords:** drones, emergency, medical care

## Abstract

**Background:** The application of drones in healthcare is a new concept being introduced in various countries to fly medical supplies. Public perceptions have been investigated but the perceptions of healthcare professionals on the frontline have not been explored. This study examined the perceptions of emergency care providers in England and Scotland to determine how they believe drones could be used when providing emergency care.

**Methods:** Frontline healthcare professionals were surveyed regarding their perceptions of drones in emergency medical scenarios. A survey with 27 questions built on SNAP 11 was published on social media for participant recruitment.

**Results:** Thematic analysis yielded insights into what healthcare professionals believe should be flown in an emergency notably: blood, defibrillators, and medication. Drones are perceived to be beneficial for life-threatening scenarios (high-risk, time-critical, trauma, search, and rescue applications) and routine medical care such as delivery of medical supplies and minor interventions. 100% of participants believed that providing medical care could benefit from a drone flying to a remote area or directly to a patient. 76% believed that having a drone fly key medical equipment faster could change the outcome of a patient. Scepticism regarding regulations and logistics involved, as well as concern for drone-based medical care, were identified by participants.

**Conclusion:** Drone use in healthcare remains an immature field and this study confirms that this domain warrants further research. It is key to remember that the perspectives of those impacted by the integration of drones will have to be explored to guide the application.

## Introduction

A drone, or unmanned aerial vehicle (UAV), is an aircraft without a pilot, guided remotely(1). The first known UAV use was in 1849, as a violent act by Austria against Venice(2). Drones are commonly used in warfare, with the USA deploying them in Afghanistan, and Ukraine using them against Russia(3,4). Other applications for drones include surveillance, photography, mapping, locating survivors and delivering supplies(5). Drones are attractive because they can bypass the constraints and limitations of road transportation with various payloads, speeds and heights(5).

The novel application of drones in healthcare has thus far included flying medical supplies such as vaccines, blood, prescription medicines, specimens and insulin in Africa, Europe, and USA(5,6).

Covid-19 accelerated the usage of drones in the healthcare field, adapting UAVs to the challenges experienced in lockdowns and maximised delivering testing samples, personal protective equipment, and vaccines(7,2).

While drones remain in their infancy stages within healthcare, UAVs are becoming increasingly appealing for emergency care settings.

Drones have successfully flown defibrillators to emergencies requiring immediate medical care(2). In 2022, a man was saved in Sweden because a defibrillator was delivered by drone before the ambulance arrived(8).

Investigations into flying organs by drone to decrease transplantation waiting times have found successful deliveries of kidney and lungs (9). Lungs were delivered in 2021 in Toronto for transplantation(10).

One study was conducted in the United Kingdom (UK) investigating flying a medical supply for an emergency case scenario(11). The study in Wales flew a defibrillator by drone in a successful simulation in 2021(11). This prompted further research into drone development in healthcare in the UK (11).

Public perception of drones is key to integrating drones into healthcare(12). Only one study considered healthcare professionals’ acceptance of drones: examining what surgeons thought about drones being used in the American organ transplantation context(13).

The current study targeted the requirement for further research on drones in healthcare. If drones are to be introduced to the UK healthcare system, it is essential to assess perceived needs. Public perceptions have been studied and trials have begun for operating drones; investigating perceptions of what a drone can carry have not. This study explored how drones could benefit medical emergencies in the UK through surveying emergency healthcare providers to determine their needs and how they could be assisted by drones in emergency situations.

## Methods

The research question for the study was “how can drones be used in emergency care settings in the United Kingdom?”.

This study explored potential uses and needs for drones in the UK when caring for a patient in an emergency medical setting by:

1. Administering a survey to investigate the perceptions of emergency healthcare professionals working on the frontline.
2. Identifying a list of items that emergency care providers believe would be beneficial to have flown in a medical emergency.
3. Analysing the data collected to create recommendations for future drone development in healthcare.
4. Informing future research for providing healthcare in the UK.

The survey was built on SNAP 11(14). It consisted of 60 questions, including multiple choice questions, open-ended questions and six medical scenarios where participants were invited to comment on what they would like a drone to carry to provide care to the patient.

The survey was distributed on social media platforms as they are a free recruitment tool and a proven recruitment method(15). The research team posted on Twitter, LinkedIn, and Facebook, for convenience sampling recruitment. Key contacts in the industry shared the link on their platforms. Email was used to contact eligible charities and organisations that provide frontline medical care. This survey was distributed throughout the UK, seeking representation from each constituent country.

Eligible participants (filtered via eligibility questions in the survey) were UK healthcare providers over the age of 18. Consent was obtained electronically at the beginning of the questionnaire, following an information landing page. Participants remained anonymous with the only personal information being their profession, organisation, and country.

Ethical approval was received by the School Ethics Review Board (SERB) of the University of Aberdeen for the Faculty Medicine, Medical Sciences and Nutrition, reference 2367.

It took six days until the recruitment of the first participant, despite analytics demonstrating high engagement with the posts and clicks on the survey link. Feedback was received that the survey was too long, and an amendment was submitted to SERB to remove 33 questions. The decision as to which questions to keep or remove was based on the three responses received. Among the 33 questions, three scenarios were removed. Approval was received and the revised shorter survey was launched.

The quantitative data was analysed using descriptive statistics in percentages. The qualitative data was analysed through Braun & Clarke’s thematic analysis methodology(16). The six steps were applied to identify, extract and present the relevant themes provided by the participants in the open-ended questions and the medical scenarios(16).

## Results

SNAP 11 does not provide analytics, so this was monitored regularly by noting the number of viewers on the link when logging into SNAP 11. As only the viewers on the link at the time of checking were noted, it is possible there were more than 207 viewers. There was a total of 17 participants, with an additional nine who began the survey but were not eligible based on their response to the first question (asking whether they are an emergency healthcare worker). While Wales and Northern Ireland were included in the recruiting strategy, no participants were yielded. This research comprises only of England and Scotland.

The participants were given a list of professions to select from and if they did not find their profession, they could tick the box “other” and complete the box with their profession.

There was a mix of geographical locations from the respondents of urban, rural, or replying “other” to include a mixture of urban and rural based on where they were sent to provide care.

When asked whether a drone could facilitate providing emergency care if it could fly to a laboratory, 65% replied “yes” and 35% replied “no”.

When asked whether a drone could enable emergency care by flying to a remote location or to an endangered patient, 100% replied “yes”.

When asked in an open-ended question whether they believed having key medical equipment brought to them faster during an emergency medical case could change the outcome of a patient, 76% replied yes. Within the other responses, a “yes” was written accompanied by something else to provide some form of scepticism or concern such as:

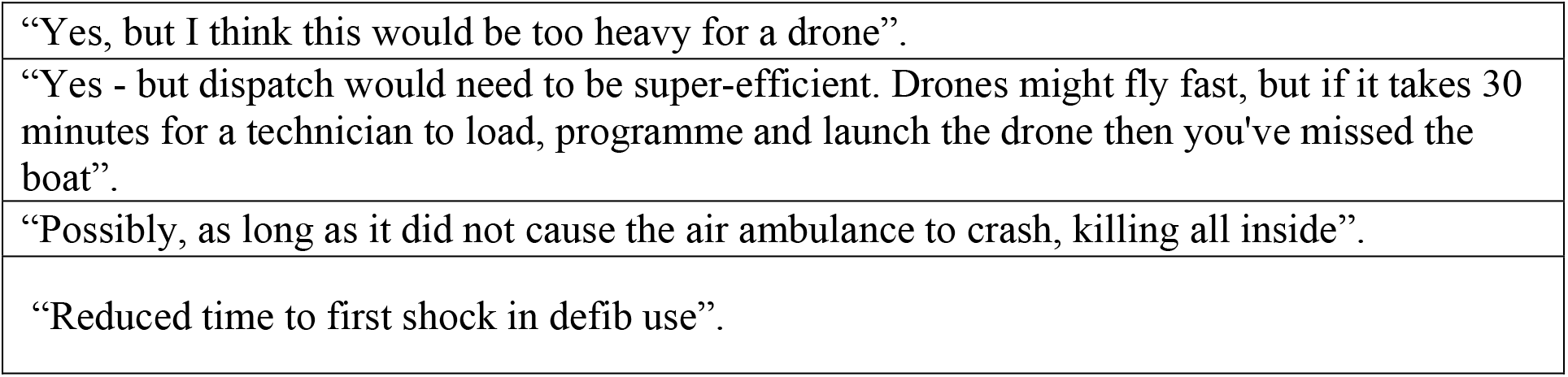

Participants were asked a series of questions regarding concerns or scepticism they may have towards the usage of drones and were invited to elaborate. Table 3 captures what was shared regarding potential obstacles or fears associated with using drones. Many of the issues raised involve establishing drones in the healthcare system such as airspace concerns and related expenses.

**Table 1:** Survey Overview Numbers

|  |  |
| --- | --- |
| Clicks on link | 207 |
| Respondents | 17 |
| England | 11 |
| Scotland | 6 |
| Wales | 0 |
| Northern Ireland | 0 |

**Table 2:** The medical professions of the participants involved in the study

| **Profession** | **Number of participants** |
| --- | --- |
| Medic | 6 |
| General Practitioner | 2 |
| Paramedic | 2 |
| Volunteer | 1 |
| Other: Doctor | 3 |
| Other: Care Assistant | 1 |
| Other: Medical Student | 1 |
| Other: Biomedical Scientist | 1 |
| Total | 17 |

**Table 3:**
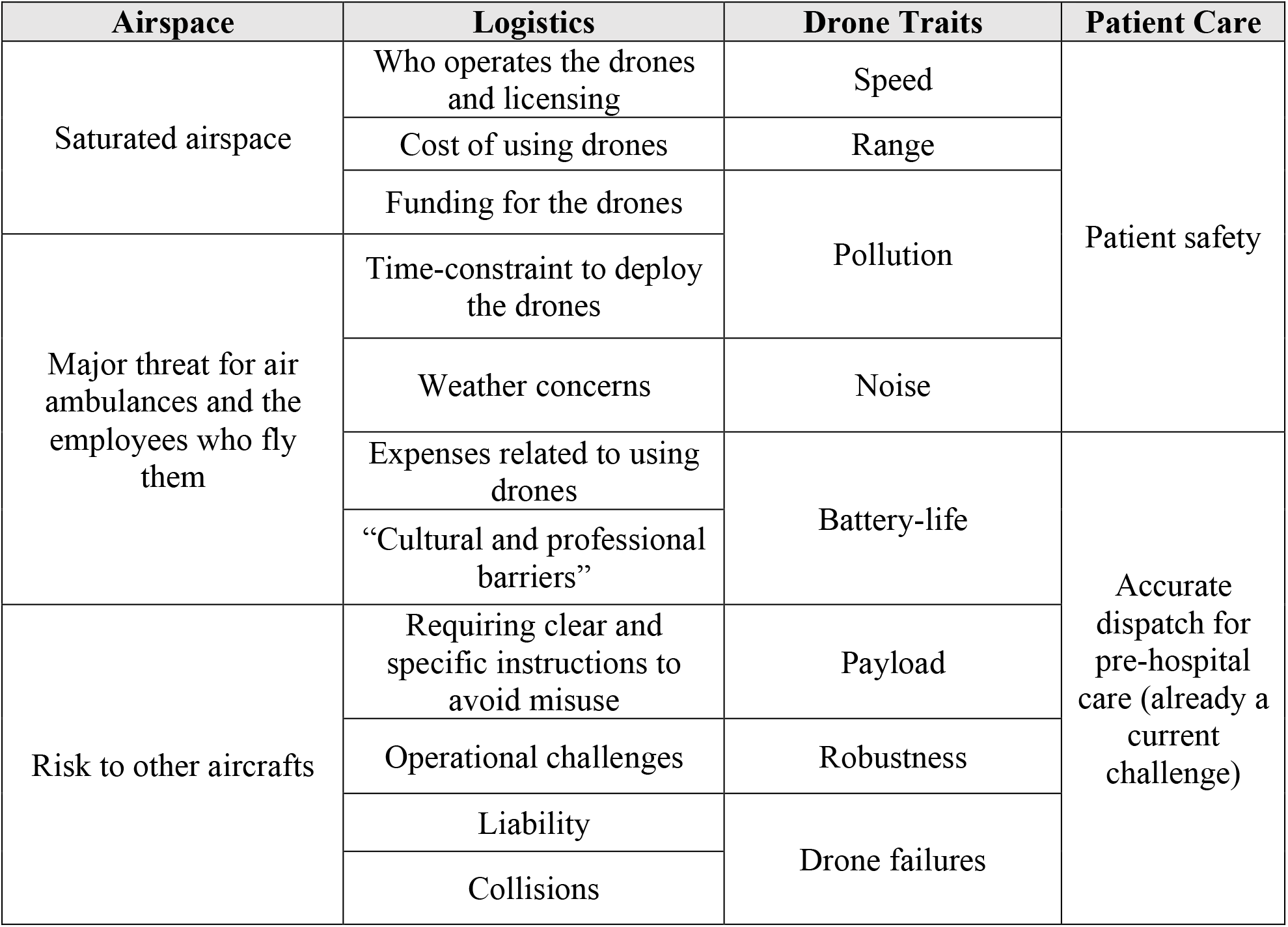
Participant concerns & scepticism related to using drones in healthcare

The participants were asked to select as many options as they wished from a list. Defibrillators and medication were the most popular responses with trauma-specific equipment and first aid kits coming in third. “Other” included blood products, vaccines, intubation kit, warming/shelter equipment, and small investigations such as an electrocardiogram.

Figure 2 shows the themes extracted from the survey and addresses the medical emergency nature that drone-based care might provide. The umbrella term “life-threatening” is divided into four case categories: high-risk, time-critical, trauma, and search and rescue. Within each category, participants provide a medical condition paired with the appropriate medical equipment to be flown. A trauma patient entrapped with severe blood loss can have the drone fly trauma specialised equipment and blood products to the team to keep the patient alive until transport to a hospital. While a patient experiencing an overdose has a window of time to reach the hospital, a drone could fly active charcoal faster.

**Figure 1:**
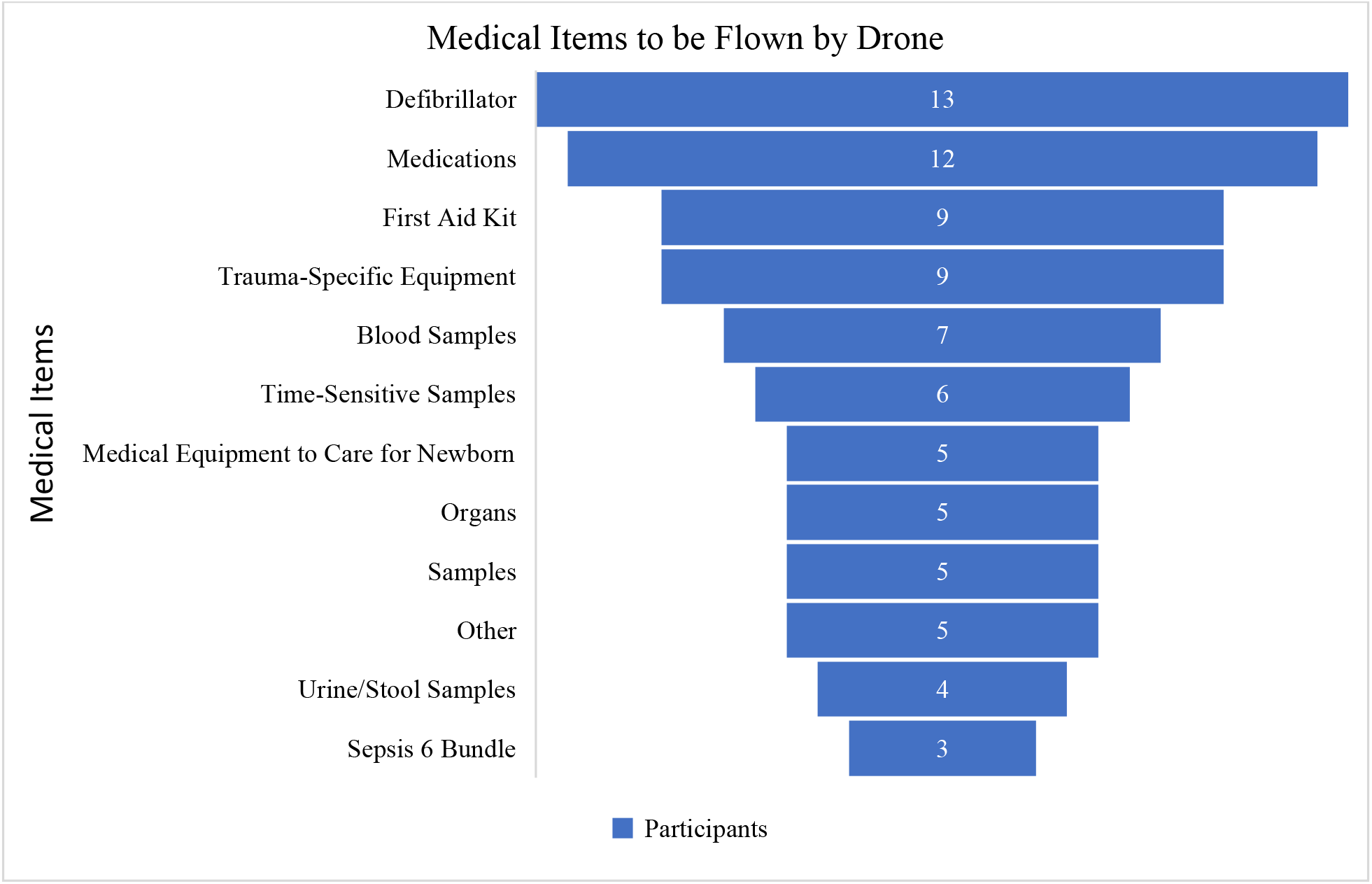
What participants would like a drone to fly in a medical emergency

**Figure 2:**
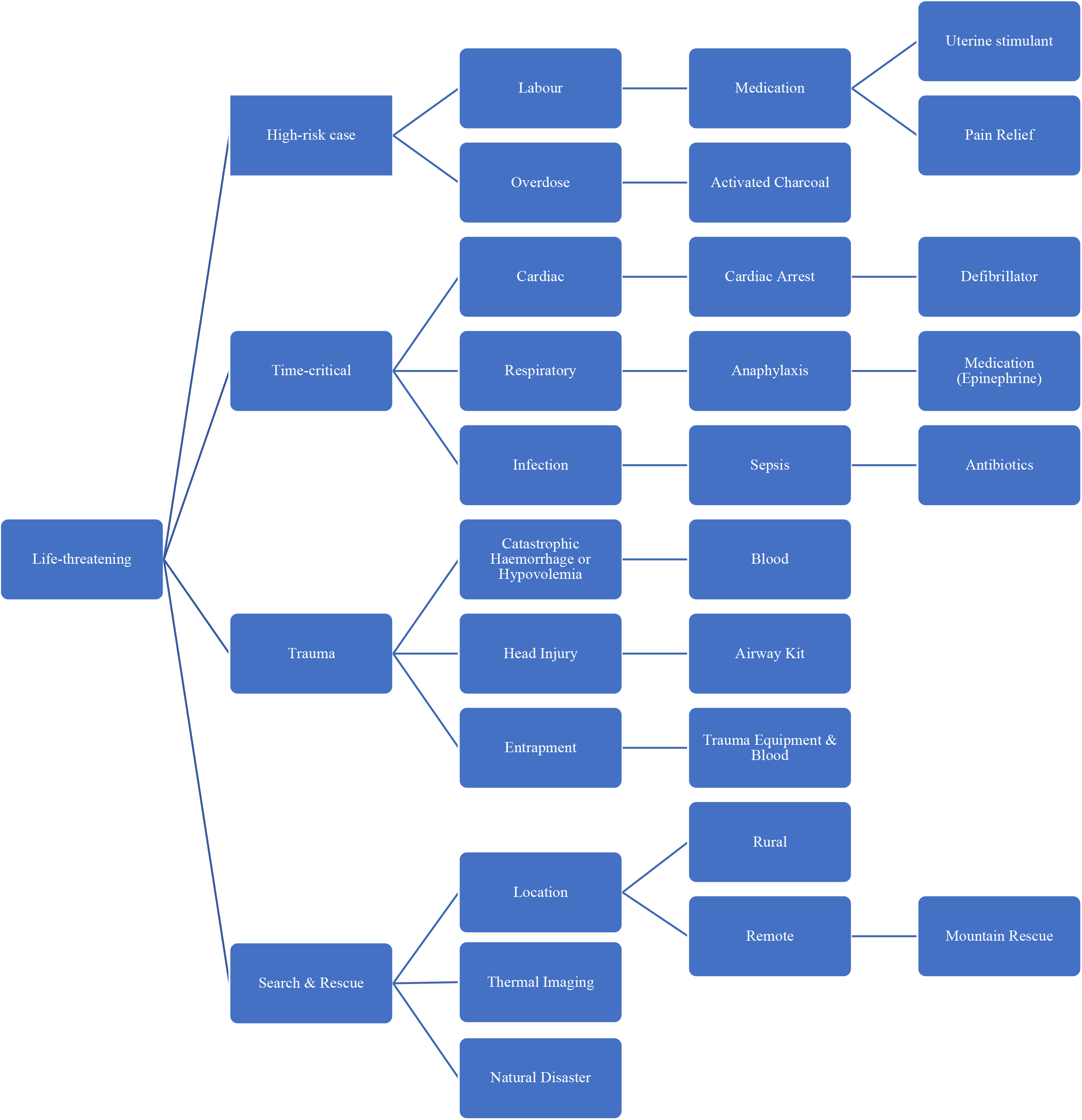
Flowchart for Pre-Hospital Emergency Medical Cases

Figure 3 presents the second theme identified: “routine care”. This was extracted from participant responses providing examples of use-cases when they believed a drone could be beneficial, falling under “standard medical care”. A drone to fly to and from a laboratory to analyse samples whether as part of a routine rural GP delivery, or an emergency would be valuable. Small interventions, specifically for rural areas that do not have access to specialised equipment at all times, could be flown by drone to aid these healthcare providers. The delivery of supplies includes vaccines to a rural practice, resupplying medical equipment that has been used in an emergency or providing additional support like a warming blanket.

**Figure 3:**
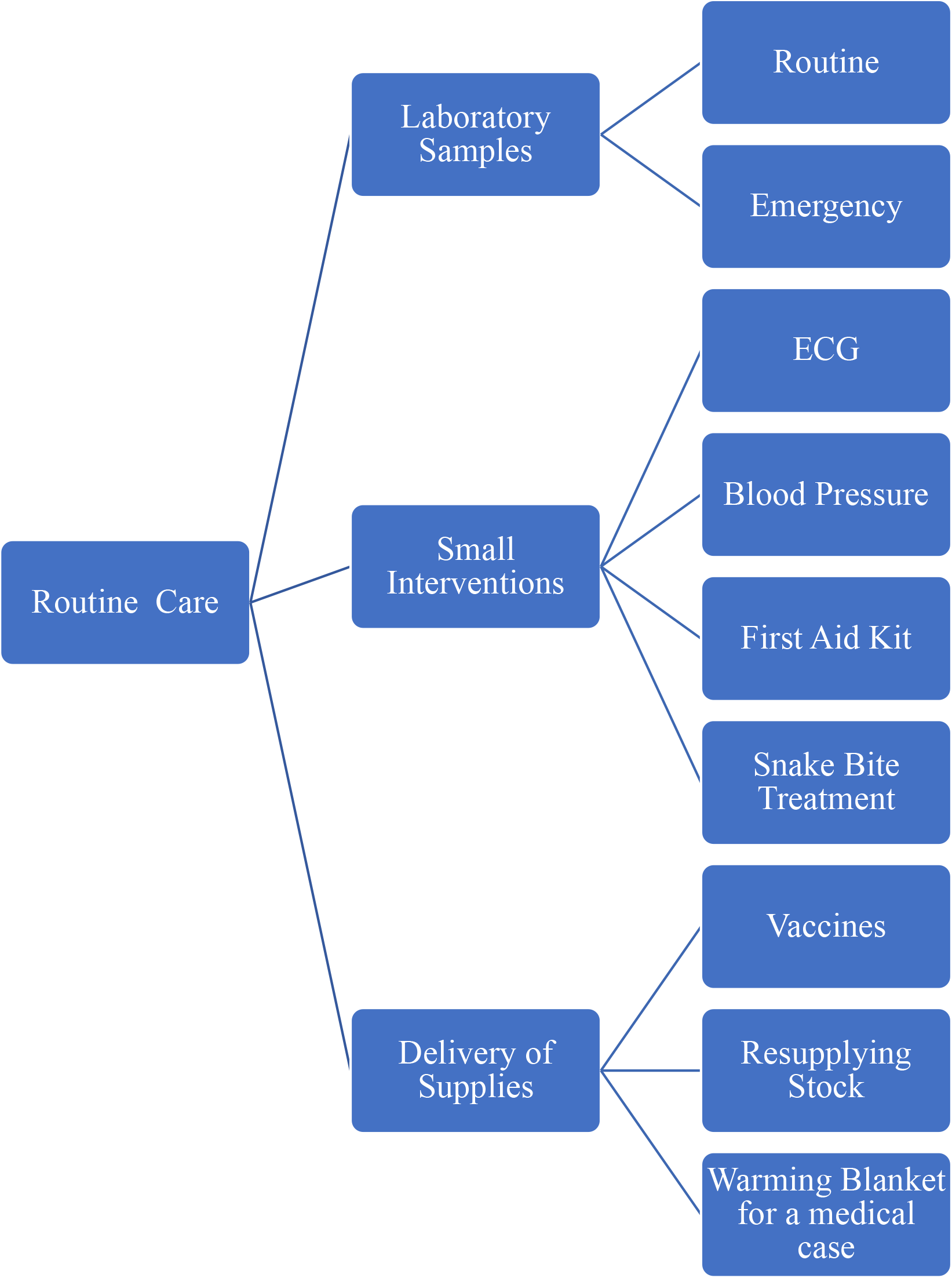
Flowchart for Application of Drones in Pre-Hospital Standard Care

70% of participants responded their desired primary medical supply being flown would be blood or blood products. Additional medical supplies were identified such as a tourniquet, dressings, and a tranexamic acid (TXA) autoinjector. Three participants wrote they could not provide an answer, due to the scenario being beyond their scope of practice.

A defibrillator was the primary medical supply identified in Figure 5. Other medical equipment reported to accompany the usage of the defibrillator were an airway kit, medication, and a LUCAS^*^.

**Figure 4:**
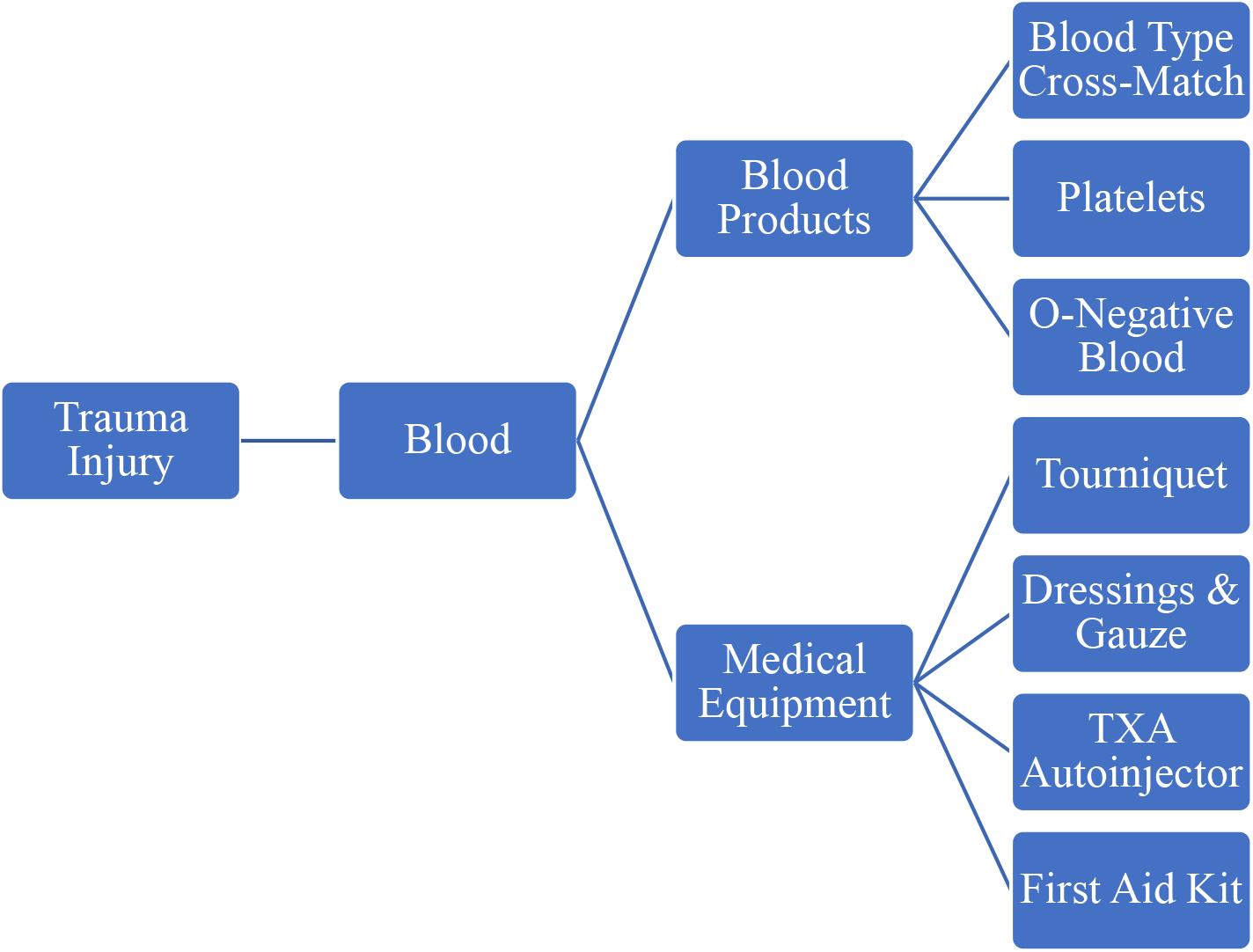
Flowchart of Scenario 1: Involving a trauma injury with severe blood loss

**Figure 5:**
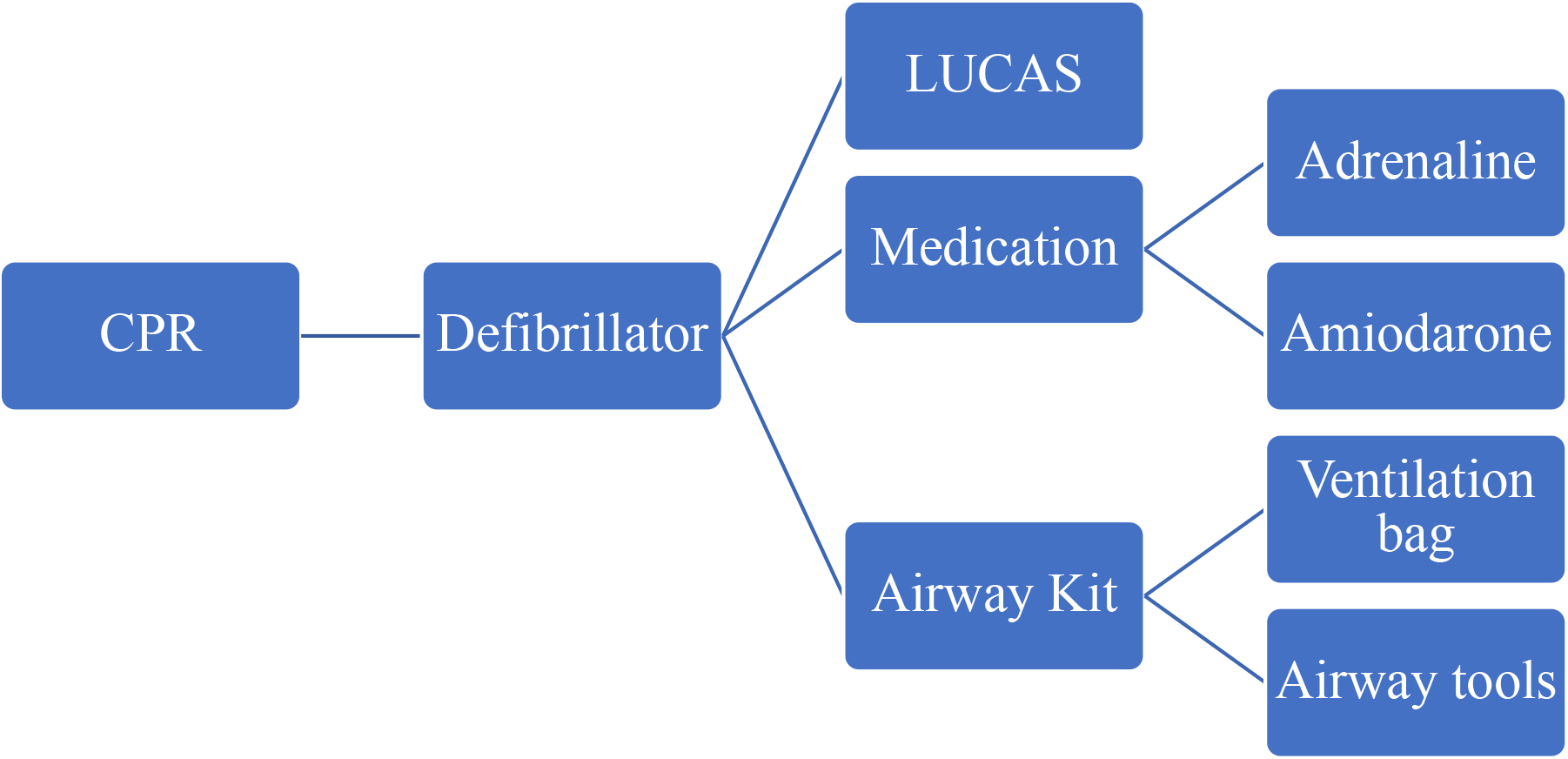
Flowchart of Scenario 2: Involving a patient requiring CPR

**Figure 6:**
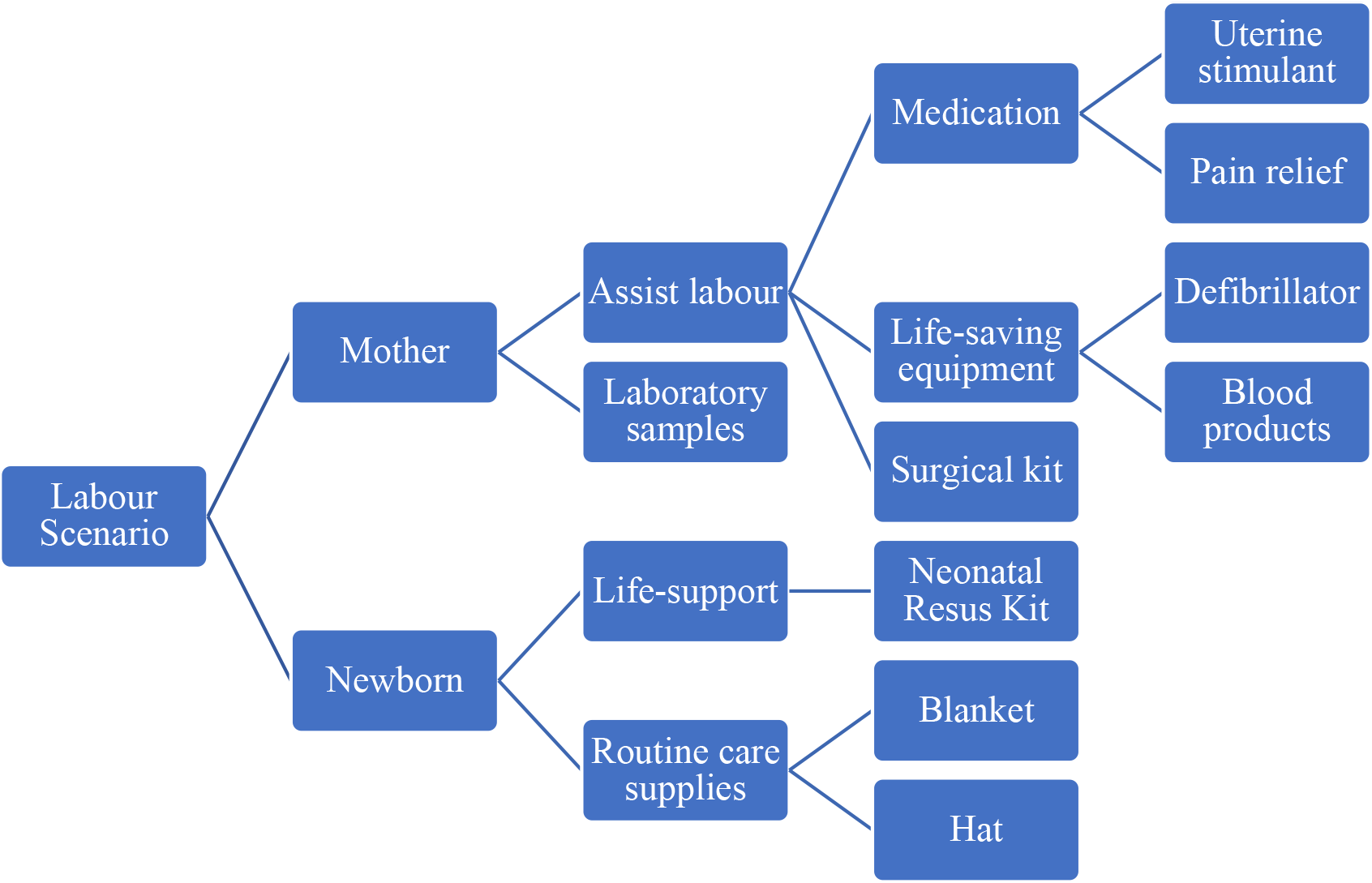
Scenario 3: Mother experiencing labour difficulties

In the scenario of labour difficulties, participants were divided, with 53% providing an example of a medical supply the drone could fly. 47% thought that they could not envision using a drone in this scenario, that the mother would need a hospital immediately, or that their rural location could not provide appropriate care, ultimately rendering medical care by drone insufficient. A participant wrote: “frankly I’d rather have a helicopter take the mother and baby elsewhere asap (sic: as soon as possible)”.

The 53% offering ideas of medical equipment, ranged from life-saving supplies like blood or a resus kit to care supplies such as pain relief and a blanket for either the mother or new-born.

## Discussion

The themes identified, life-threatening and routine care, both harness the capabilities a drone can offer such as flying rapidly to a medical emergency and reaching rural locations requiring medical supplies.

The main themes correspond with the three main medical supplies identified: blood, defibrillator, and medication. Blood coincides with trauma cases such as hypovolemia or catastrophic haemorrhage. Flying essential medication to provide care to a patient experiencing a life-threatening emergency can vary based on the setting. Anaphylaxis was an example provided by participants where adrenaline could be flown to care for this patient. The participants reported other medical supplies, but these were in response to specific questions or prompts such as snake bites.

For a traumatic injury, the consensus was that a drone could deliver blood products. Participants did note that based on their specific profession they may have blood with them but could be resupplied by a larger hospital via drone.

Blood is not listed in Figure 1 but was written explicitly under “other” for participants and is evidently something that would be valuable to have flown.

Regarding the CPR scenario, participants primarily reported a defibrillator and a LUCAS; this was specified to rural or remote cases for some participants. Participants did note that they usually have a defibrillator on them so this could be for bystander to receive a defibrillator if one wasn’t available. Given that drones can broaden the possibilities for providing medical care, the role of the bystander would need to be evaluated and taken into consideration. For other questions asking what drones could carry, a participant wrote: “AED (defibrillator) for bystander performing

CPR”. This is supported by the literature where bystanders using defibrillators can improve survival rate(17).

The mother in labour scenario is the only one that does not describe the medical care the patient requires, only describing “labour difficulties” to invite creative thinking, interpretation and address the unplanned nature of childbirth. Responses showed a divide between participants. Some who did provide an answer for a medical supply expressed discomfort with drone-based care. Using drones for childbirth care is unprecedented, however, a novel pilot project in Botswana is addressing the maternal mortality associated with childbirth by delivering blood or medication to four villages to provide life-saving care to pregnant women(18).

A participant submitted a compelling response: “I think your focus is about what it can carry - perhaps think more broadly about what it can do e.g., ‘eyes’ to search for casualty, thermal imaging at night, etc.”. It is possible that researchers are limiting themselves by focusing solely on what a drone can carry.

Integrating drone-based medical care into the British healthcare system would involve logistics on multiple levels. This would include licensing for flying drones, responsibility for dispatching drones, establishing relevant contexts, what the drone is containing and the direction of flight for the drone (i.e., to or from a patient).

The drone-based care team would be extensive based on the dispatch and receival of drones, not solely the team present with the patient. Additionally, current healthcare providers cannot spend time while caring for a patient to load and fly the drone, anything they do must be very rapid.

A participant responded: “Although ED is so busy now if a responsive service with drones could beat the ED queue?”. This is a fascinating idea to consider and contemplate the changes to be made if drones were fully integrated, broadening the implication of drone-based care.

Findings that blood is a valuable medical item to fly within done-based care supports current literature. Rwanda integrated blood delivery with drones in 2016 to improve blood delivery and care(19). 43% of drone deliveries, from 2017 to 2019 in Rwanda were emergency blood deliveries and had quicker dispatches than ground transportation(19). A study in Montreal simulated the delivery of blood products from a blood bank to a hospital, comparing drones with ground transportation, finding that drones were faster and offered significant time benefits, which would be beneficial during a mass casualty incident(20). Additionally, Japan flew blood in drones in a transoceanic flight to reach remote islands to provide blood transfusions(21).

The factor predicting survival for out-of-hospital cardiac arrest is the response time(22). Boutilier et al modelled that drones could fly and deliver a defibrillator faster than the 911 response median time in Toronto (23). A Swedish pilot-study investigated flying a drone for real-life cardiac emergencies and established that defibrillators were successfully dispatched and received in 92% of cases(24). Regarding the integration of drones, a study in North Carolina found that a theoretical drone network to deploy 500 drones to cardiac arrest cases would double expected survival rates due to rapid response while being cost-effective(25). These studies correspond with the current study participants reporting a need to fly defibrillators.

Adrenaline and antivenom for snake bites have been tested for drone flight and found that the integrity was not changed(26,27). Antibiotics have been considered a good candidate for flight to treat sepsis(27). Further research should be conducted for flying medications by drone and other medical supplies like the LUCAS have not been investigated.

Table 3 presents valid, self-explanatory concerns noting logistical aspects that would need to be organised prior to integrating drones into healthcare and developing appropriate regulations. Cost is written as “the potential being outweighed by the expenses” by a participant. There was negativity surrounding the usage for drones where “ambition is ahead of capability”. Collisions were reported as a concern, with an air ambulance professional writing: “drones are one of the biggest risks to my life in the air”. While drone crashes are rare, regulations must be developed to address the event of a drone crash (29). In comparison, there was enthusiasm expressed as: “No, think they could be revolutionary”.

Ghana and Rwanda have harnessed medical drones to reduce their carbon footprint(30). Drones emit low carbon emissions because they run on batteries instead of fuel, making them friendlier for the environment rendering the “pollution” concern in Table 3 inapplicable(30).

Natural disasters were reported without further elaboration. Drones have been used in post-natural disaster settings to provide humanitarian aid by delivering supplies to remote areas or searching for survivors(31).

While social media for research recruitment has its strengths, it requires a network to be useful. The student researcher is an international student and does not have an extensive network in the UK, meaning the recruitment was limited to supervisors and few key contacts in the industry.

It was not possible to verify the eligibility of participants. The ambiguity of this survey is evident through nine respondents beginning the survey while being ineligible. Selection bias is a concern through convenience sampling as it was anticipated that participants who have knowledge of drones or an interest/dislike towards drones would be participating. Future studies should include a larger sample size.

The study faced time constraints and as such, only SERB was contacted, rather than also going through NHS ethics. Time constraints were addressed by keeping the survey open for as long as possible.

The survey struggled with a low response rate despite high engagement. It was anticipated there may be a greater sample collected from Scotland due to the University of Aberdeen and large representation from England due to population size.

Research fatigue was a major deterrent for completing the survey as two contacts denied our request to help distribute the survey due to research fatigue experienced by healthcare professionals(32). This was further confirmed by a charity responding saying they can only participate in a limited number of studies per year and had already maximised that number.

A notable strength of this study is the exploratory nature to investigate the perceptions of frontline healthcare workers who would be impacted by drone integration.

The insights offered warrant further research. This survey provides valuable insights to guide future research and drone governance. While it is possible to continue investigating what drones can fly and how to make the science behind flying medical equipment viable, it will remain just as essential to consider the perspectives of those who would be involved in integrating drones into healthcare. It would be valuable to investigate perceptions of groups individually like paramedics, nurses, or laboratory workers and then distinguish the relevant medical cases for those groups.

Regardless of drones being notorious for politics, warfare, and terrorism, the healthcare sector could utilize the abilities that drones have to offer to better the efficiency of their services(33).

## Data Availability

Data is available on request from the authors. Please contact

## Acknowledgements

We would like to thank the participants of the survey and contacts that helped with recruitment.

## Footnotes

* A LUCAS is automized medical equipment that conducts chest compressions for CPR.

## Notes

### Competing Interest Statement

Supervisor Dr. Sophie Barrack has honorary status with the University of Aberdeen and is employed by Apian, a medical drone development company.

### Funding Statement

This study did not receive any funding.

### Author Declarations

The School Ethics Review Board (SERB) of the University of Aberdeen for the Faculty Medicine, Medical Sciences and Nutrition gave ethical approval for this work. Reference 2367.

